# Unexplained Dyspnea and Cardiopulmonary Exercise Testing: Rationale for a Dyspnea Clinic

**DOI:** 10.1101/2021.11.05.21265990

**Authors:** Pooja Nair, Jeffrey W Christle, Constance Verdonk, Nicholas Cauwenberghs, Yair Blumberg, Laura S Sasportas, Roham Zamanian, Euan Ashley, Tatiana Kuznetsova, Jonathan Myers, Francois Haddad, Myriam Amsallem

## Abstract

**Objective:** To describe the experience of a tertiary-care center in the management of patients with unexplained dyspnea referred for cardiopulmonary exercise testing (CPX).

**Methods:** All consecutive adults with unexplained dyspnea who underwent CPX at a single tertiary referral center over a ten-year period were included from a prospective registry. We collected data on final diagnosis, routine labs and diagnostic testing including stress/resting echocardiography performed within 6 months of CPX.

**Results:** From 2008 to 2019, 156 patients with unexplained dyspnea were referred for CPX with no prior history of cardiovascular disease, lung disease or neuromuscular disease. A 4-fold increase in referrals was noted during this time period. Analysis of diagnostic work-up revealed marked heterogeneity, particularly according to the specialty of the referring physician. Among the 134 patients who achieved an adequate level of exertion during CPX, 24 (17.9%) and 30 (22.4%) patients had an abnormal age-predicted peak VO_2_ of < 80% according to Wasserman’s and FRIEND equations respectively. Further analysis revealed 22.5% of patients had a VE/VCO2 slope > 34, suggesting ventilatory inefficiency. Subgroup analysis of 108 patients with complete left ventricular (LV) diastology revealed 13% of patients with a final diagnosis of unexplained dyspnea after CPX were found to have underlying diastolic dysfunction.

**Conclusions:** Our study illustrates an increase in CPX referrals for unexplained dyspnea and associated heterogeneity in diagnostic testing in this population. There is a need for an integrated dyspnea clinic to standardize workflow and facilitate early diagnosis for these patients.

## INTRODUCTION

Dyspnea is the most frequent symptom of cardiopulmonary disease, albeit not specific. Patients with unexplained dyspnea are often exposed to heterogeneity in diagnostic work-up, frequently delaying the diagnosis up to 2 years [1]. Cardiopulmonary exercise testing (CPX) combines the advantages of exercise testing (i.e. provocative tests using exercise on a treadmill or bicycle with hemodynamic monitoring, ECG tracing and symptoms) with measurement of ventilatory parameters and gas exchange during progressive exercise [2]. While the diagnostic utility of CPX in patients with unexplained dyspnea has been previously reported [3], its incremental value on novel markers of early left ventricular (LV) dysfunction (such as global longitudinal strain or diastolic dysfunction at rest) is underexplored.

This study aims to report the experience of a large tertiary single-center in the management of patients with unexplained dyspnea referred for CPX over a decade. We hypothesize that novel markers of early LV dysfunction can be incremental to CPX findings to identify the etiology of dyspnea.

The first objective was to evaluate the number of patients with unexplained dyspnea referred to CPX over the years, their clinical profile and type of diagnostic tests other than CPX performed (including stress and rest echocardiography, laboratory data, pulmonary function testing (PFT) or chest X-rays (CXR) when available). The second objective was to assess the utility of CPX in determining the etiology of dyspnea. The third objective was to further investigate patients clinically determined to have “unexplained dyspnea” after clinical work-up, particularly in light of novel echocardiographic metrics of diastology to detect early LV dysfunction in the subgroup with available diastolic profile and strain data.

## METHODS

### Study population

This retrospective study used the StudyTRAX database of all exercise testing studies performed at Stanford from March 1st 2008 to February 13th 2019 to select all patients with unexplained dyspnea. Inclusion criteria were: adults (age ≥18 years) with unexplained dyspnea referred for CPX with complete stress ECG data. If several tests were performed, only the first visit was selected. Exclusion criteria were: previously known cardiovascular disease (including heart failure with reduced ejection fraction (HFrEF), heart failure with preserved ejection fraction (HFpEF), valvular heart disease, heart or heart-lung transplant, hypertrophic cardiomyopathy, ischemic cardiomyopathy, restrictive cardiomyopathy, pulmonary hypertension, congenital heart disease and/or arrhythmia such as atrial fibrillation or flutter), known lung disease (including chronic obstructive pulmonary disease, severe asthma, restrictive lung disease, history of chest radiation therapy, chronic thromboembolic disease), neuromuscular disease (including Duchenne or Becker muscular dystrophy), research studies or patients not referred for dyspnea (including chest pain, syncope or palpitations).

### Cardiopulmonary testing

CPX tests were performed on a treadmill or upright bike, using individualized RAMP protocol, with an integrated metabolic cart (Quark CPET, CosMed USA Inc., Concord, CA, USA) using breath-by-breath data capture and analysis [4]. Peak heart rate is presented as absolute value and as percentage of predicted heart rate. Peak systolic blood pressure is presented as absolute value and corrected by level of external work at peak (expressed as mmHg/METs) as previously validated [5]. The achievement of a respiratory exchange ratio (RER= VCO_2_/VO_2_) > 1.05, or between 1.00 and 1.05 with a rating of perceived exertion (6–20) of ⥸16 were used to determine peak effort. Peak VO_2_ was calculated as the highest average VO_2_ over 30 s during the last phase of exercise and presented as age-predicted values using the Wasserman/Hansen or FRIEND equations [6] (**Supplementary Table S1**). Minute ventilation (VE) and carbon dioxide output (VCO_2_) measured throughout the entire exercise bout (i.e. including every data point) were used to calculate the VE/VCO_2_ slope via least squares linear regression (VE = a VCO_2_ + b, a = slope) as previously validated [7, 8]. The partial pressure of end-tidal CO_2_ (P_ET_CO_2_), the oxygen uptake efficiency slope and O2 pulse were calculated as previously described [2]. Resting forced expiratory volume in 1 s (FEV_1_) was measured before starting CPX. The breathing reserve (BR) was expressed as the difference between the maximal voluntary ventilation (MVV; defined as FEV1 × 40) and the maximum exercise ventilation (VE).

A continuous 12-lead ECG was monitored during exercise and recovery. ST segment elevation and depression were measured by a certified-technician per international consensus criteria [9]. Ventricular or supraventricular arrhythmias during or after exercise was collected. Shortness of breath reported by the patient during exercise or recovery was noted in addition to the primary subjective symptom eliciting test termination.

### Echocardiography

Resting and/or stress echocardiograms performed within 6 months of the CPX date were included. Echocardiograms were performed using commercially available echocardiographic systems (iE33 or EPIQ 7□C; Philips Medical Imaging, Eindhoven, the Netherlands), according to the American Society of Echocardiography (ASE) guideline recommendations [10]. Image analyses were performed on Xcelera workstation by a level 3 board-certified cardiologist (MA). No patient was in atrial fibrillation at the time of echocardiography analysis. Area and volumes were indexed to body surface area. Standard echocardiographic views were obtained in two-dimensional and color tissue Doppler modes. LV internal diameter, posterior wall and interventricular wall thickness were measured in end-diastole on the parasternal long axis, enabling calculation of the LV mass and the relative wall thickness ratio, defining LV concentric hypertrophy, eccentric hypertrophy or concentric remodeling per ASE guidelines [10]. LV end-diastolic volume, end-systolic volume and LV ejection fraction were calculated using Simpson’s method. LV global longitudinal strain was obtained using Lagrangian strain by manual tracing from the apical 4-chamber view and calculated in the following formula: 100□×□(L_1_-L_0_)/L_0_ as previously validated [11]. The threshold for longitudinal LV strain was based on epidemiology-based and outcome-driven criteria with values greater than -17.4% and -18.8% in men and women respectively, classified as abnormal LV deformation [12, 13]. Transmitral pulsed-wave Doppler velocities (E, A waves) and tissue Doppler velocity (e’) of the lateral and septal mitral annulus were obtained from apical 4-chamber view, enabling determination of LV diastolic function and filling pressure estimations. LV diastolic function was classified as grade 1, 2 and 3 according to the American Society of Echocardiographic latest guidelines [14], and as mild, moderate and severe using the previously published Redfield’s classification [15]. Left atrial (LA) volumes were obtained using biplane method of disks method, LA emptying fraction was calculated from the maximal and minimal LA volume, and LA reservoir strain was calculated using Lagrangian strain. Right ventricular size was assessed based on basal and mid-levels end-diastole diameters on the apical 4-chamber view, while right ventricular function was assessed based on the tricuspid annular plane systolic excursion. Right ventricular systolic pressure was estimated from the sum of the tricuspid regurgitation maximal velocity using the modified Bernoulli equation and estimated right atrial pressure. This latter was estimated as 3□mmHg if the inferior vena cava diameter ≤2.1□cm and collapsed >50%, 15□mmHg if the diameter >2.1□cm and collapsed <50%, and 8□mmHg in scenarios in which the diameter is enlarged or collapse index is sub-optimal otherwise. Stress-induced presence of wall motion abnormalities and right ventricular systolic pressure at peak exercise (or early recovery for patients who underwent treadmill exercise testing) or late recovery was collected when available.

### Clinical, functional and hemodynamic data

Hypertension was defined as previously diagnosed hypertension, treated or not. Dyslipidemia was defined as hypercholesterolemia previously diagnosed, treated or not. Diabetes mellitus was defined as previously diagnosed (on antidiabetic therapy or not) or when HbA1c was > 6.5%. Routine lab analyses available within 3 months before or after stress testing were collected, as well as CXR and PFT results within 12 months before or after stress testing.

### Outcomes

The final clinical diagnosis was collected via chart review of follow-up notes, which was available in 125 patients. Although Cardiology and Pulmonology follow-up notes were prioritized, all notes from other specialties were also reviewed for additional information and the Community View feature on Epic was utilized to review outside records. Follow-up was concluded in June 2020, and death was verified through chart review and follow-up.

### Statistical analysis

Data is presented as median and interquartile range for continuous variables and as number and percentage for categorical variables, and compared using Mann-Whitney and Chi-square tests respectively. P-values < 0.05 were considered statistically significant. Statistical analysis was performed using SPSS statistical software (SPSS V.26, Inc, Chicago, IL).

## RESULTS

### Study population

A total of 156 patients referred to CPX for unexplained dyspnea were included (**Figure 1A**). Over the study period, there was a 3-fold increase in the number of patients referred each year (**Figure 1B**). Baseline characteristics of the study population are presented in **Table 1**. Seventeen percent of patients had at least two of three cardiovascular risk factors (hypertension, dyslipidemia or diabetes) and 4% had all three risk factors. Over half the cohort was overweight or obese. PFTs indicated that 30% of the cohort had underlying lung disease, of which 23% was classified as mild. **Figure 1C-D** illustrates testing heterogeneity, particularly according to referring physician specialty, with only 15% of patients found to have an NT-proBNP result available within 3 months of CPX.

**Table 1.** Characteristics of the total study cohort with unexplained dyspnea at the time of exercise testing.

| **Baseline Characteristics** | **Total cohort**<br>**n=156** |
| --- | --- |
| **Age (years)** | 49 [37; 64] |
| **Male Gender, *n* (%)** | 66 (42.3%) |
| **Cardiovascular Risk Factors** |  |
| Males > 50 years, *n* (%) | 37 (23.7%) |
| Females > 60 years, *n* (%) | 25 (16.0%) |
| Hypertension, *n* (%) | 44 (28.2%) |
| Dyslipidemia, *n* (%) | 43 (27.6%) |
| Diabetes mellitus, *n* (%) | 11 (7.1%) |
| Presence of two out of three risk factors (hypertension, dyslipidemia or diabetes) | 26 (16.7%) |
| Presence of all three risk factors (hypertension, dyslipidemia or diabetes) | 6 (3.8%) |
| Smoking history, *n* (%) | 39 (25%) |
| Active smoker or quit in the last 3 years, *n* (%) | 7 (4.5%) |
| Family history of cardiovascular disease *n* (%) | 10 (6.4%) |
| Body mass index (WHO Classification), median [range] | 26 [23; 29] |
| Underweight & Normal, *n* (%) | 68 (43.6%) |
| Pre-obese, *n* (%) | 53 (34.0%) |
| Obese Class I, *n* (%) | 18 (11.5%) |
| Obese Class II, *n* (%) | 14 (9.0%) |
| Obese Class III, *n* (%) | 3 (1.9%) |
| **Pre-existing Medical Conditions** |  |
| Childhood asthma or suspected asthma, *n* (%) | 22 (14.1%) |
| Gastroesophageal reflux disease, *n* (%) | 22 (14.1%) |
| Previous cancer, *n* (%) | 11 (7.1%) |
| Obstructive sleep apnea, *n* (%) | 27 (17.3%) |
| History of thromboembolism (DVT or PE), *n* (%) | 4 (2.6%) |
| Peripheral vasculitis, *n* (%) | 2 (1.3%) |
| **Current medications (n = 147)** |  |
| Antihypertensives, *n* (%) | 32 (21.8%) |
| Antidiabetics, *n* (%) | 9 (6.1%) |
| Statin, *n* (%) | 28 (19.0%) |
| Antidepressants, *n* (%) | 28 (19.0%) |
| Antipsychotics, *n* (%) | 3 (2.0%) |
| Antireflux therapy, *n* (%) | 37 (25.2%) |
| All inhaler therapy, *n* (%) | 36 (24.5%) |
| Steroid inhaler therapy, *n* (%) | 26 (17.7%) |
| Antiarrhythmics, *n* (%) | 4 (2.7%) |
| Antianginal therapy, *n* (%) | 5 (3.4%) |
| Antiepileptics, *n* (%) | 5 (3.4%) |
| Methotrexate, *n* (%) | 2 (1.4%) |
| Aspirin, *n* (%) | 30 (20.4%) |
| Beta blocker, *n* (%) | 27 (18.4%) |
| Thyroxine, *n* (%) | 16 (10.9%) |
| **Pulmonary Function Testing (n=69)** |  |
| Obstructive lung disease | 12 (17.4%) |
| Mild | 8 (11.6%) |
| Moderate | 2 (2.9%) |
| Severe | 1 (1.4%) |
| Positive methacholine test | 1 (1.4%) |
| Restrictive lung disease | 10 (14.5%) |
| Mild | 8 (11.6%) |
| Moderate | 1 (1.4%) |
| Severe | 1 (1.4%) |
| Others: decreased DLCO | 5 (7.2%) |
| **Chest X-rays (n=63)** |  |
| Hyperinflation or emphysema | 5 (7.9%) |
| Kyphosis, moderate scoliosis or pectus excavatum | 4 (6.3%) |
| Mild atelectasis | 4 (6.3%) |
| Signs of pulmonary edema | 2 (3.2%) |
| Mild pleural effusion | 1 (1.6%) |
| Paralyzed hemidiaphragm | 1 (1.6%) |
| **Pulse Oximetry (n = 113)** |  |
| Pulse Oximetry, median [range]* | 98 [97; 99] |
| **Laboratory Data** |  |
| Hemoglobin (g/dL), median [range] (n=79) | 13.6 [12.8; 14.5] |
| Abnormal hemoglobin, n (%) | 9 (11.5%) |
| Hemoglobin < 10g/dL, n (%) | 0 |
| Thyroid stimulating hormone (mIU/L), median [range] (n=51) | 1.8 [1.1; 2.3] |
| Abnormal thyroid stimulating hormone, n (%) | 3 (5.9%) |
| NT-proBNP (pg/mL), median [range] (n=24) | 47 [30; 69] |
| NT-proBNP > 300 pg/mL, n (%) | 2 (8.3%) |
| NT-proBNP > 500 pg/mL, n (%) | 1 (4.2%) |
| Glomerular filtration rate (mL/min/1.732 m<sup>2</sup>), median [range] (n=80) | 85.5 [72; 106] |
| Glomerular filtration rate > 60 mL/min/1.732 m<sup>2</sup>, n (%) | 70 (87.3%) |
| Glomerular filtration rate 30 - 60 mL/min/1.732 m<sup>2</sup>, n (%) | 7 (8.9%) |
| Glomerular filtration rate < or = 30 mL/min/1.732 m<sup>2</sup>, n (%) | 2 (2.5%) |
| **Arterial blood gas (pH, pO<sub>2</sub>, pCO<sub>2</sub>) (n=10)** |  |
| pH, median [range] | 7.42 [7.40; 7.45] |
| pO<sub>2</sub>, median [range] | 89.3 [80.6; 111.5] |
| pCO<sub>2</sub>, median [range] | 40.6 [34.4; 45.0] |

Data is presented as median [interquartile range] or number (percentage). DVT: deep venous thrombosis; PE: pulmonary embolism. \*No patient had oxygen saturation level < 92%.

**Figure 1.**
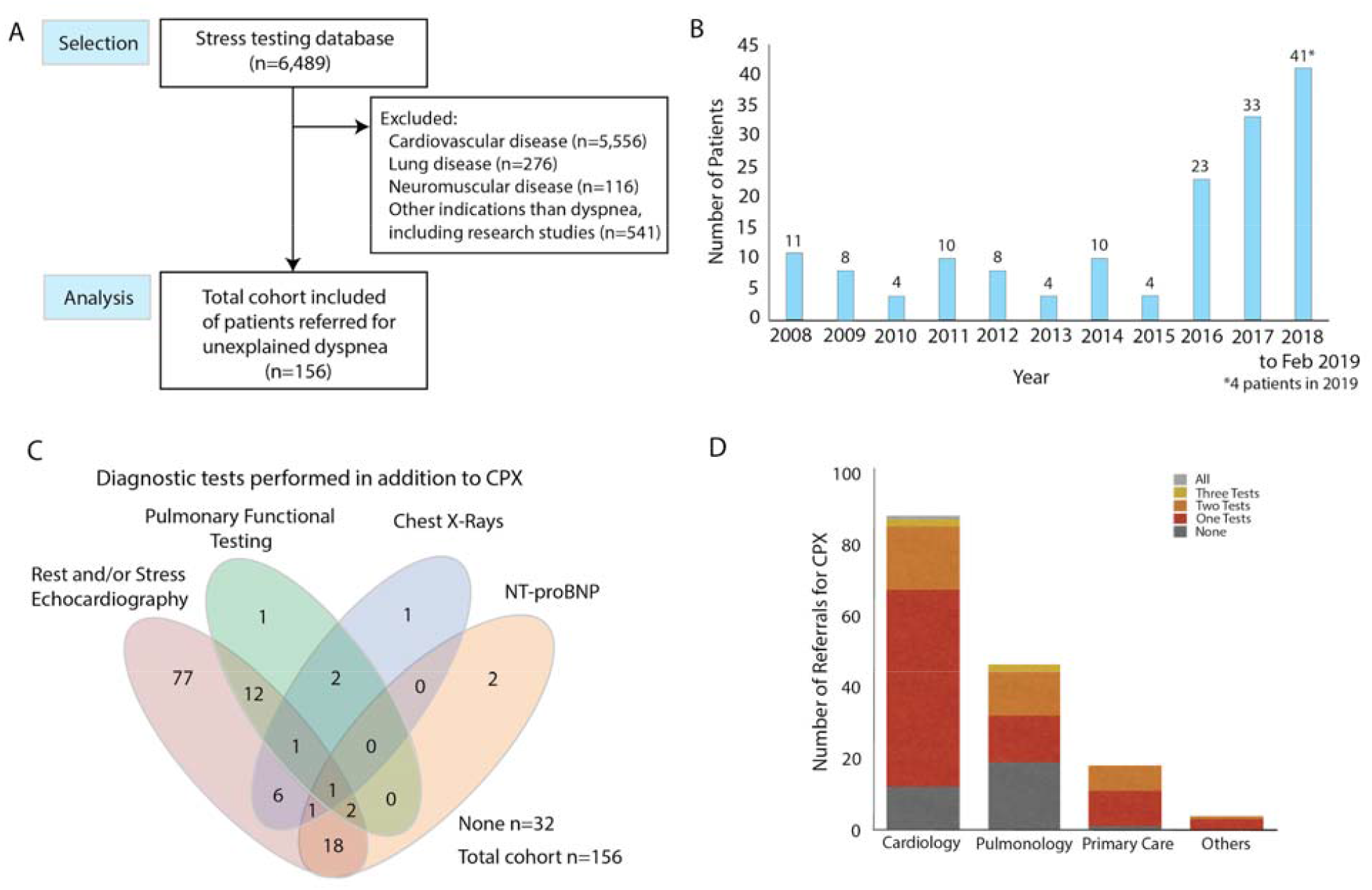
Flow chart of the study population (A) and number of patients with unexplained dyspnea referred to CPX per year (B). Heterogeneity of the tests performed in addition to CPX (C) and availability of ancillary testing prior to CPX based on the specialty of the referring physician (D). *PFT/Chest X-Rays within 12 months, echocardiogram (at Stanford Health Care Hospitals or outside of Stanford Health Care Hospitals) within 6 months and NT-proBNP within 3 months of CPX.

### CPX findings

CPX results for the total cohort are presented in **Table 2, Figure 2A** and **Figure 2B**. About half of the patients reported shortness of breath at peak exercise. Thirty-three patients were not able to reach an RER > 1.05 consistent with deconditioning or other functional limitations. Within this group, 11 patients reached subjective maximal effort (defined by an RER <1.00 or between 1.00 - 1.05 with a perceived exertion of 16 or more. Twenty-two patients had an RER < 1.00 or between 1.00 - 1.05 with a perceived exertion < 15, suggestive of undermotivation or apprehension that limited achievement of physiological peak exercise performance. Reasons for stopping in this group included discomfort with the mouthpiece, generalized fatigue, dyspnea, leg pain and increasing ventricular ectopy. Peak heart rate > 85% of predicted was achieved in 73.1% of patients, significantly more in patients without beta blockers (76.7% versus 55.6% in patients with beta blockers, p=0.03). In 102 patients, the VE/VCO2 slope was available for re-analysis of the two VT1 and VT2 slopes. While the clinical report indicated 23/102 (22.5%) with a slope >34, the VT1 slope was > 34 in 8/102 (7.8%) patients, and the VT2 slope was > 34 in 15/102 (14.7%) patients. **Figure 2B** illustrates the proportion of patients in each primary CPX variable group based on the 2016 guidelines, besides PETCO2 which was unavailable for most patients [3].

**Table 2.**
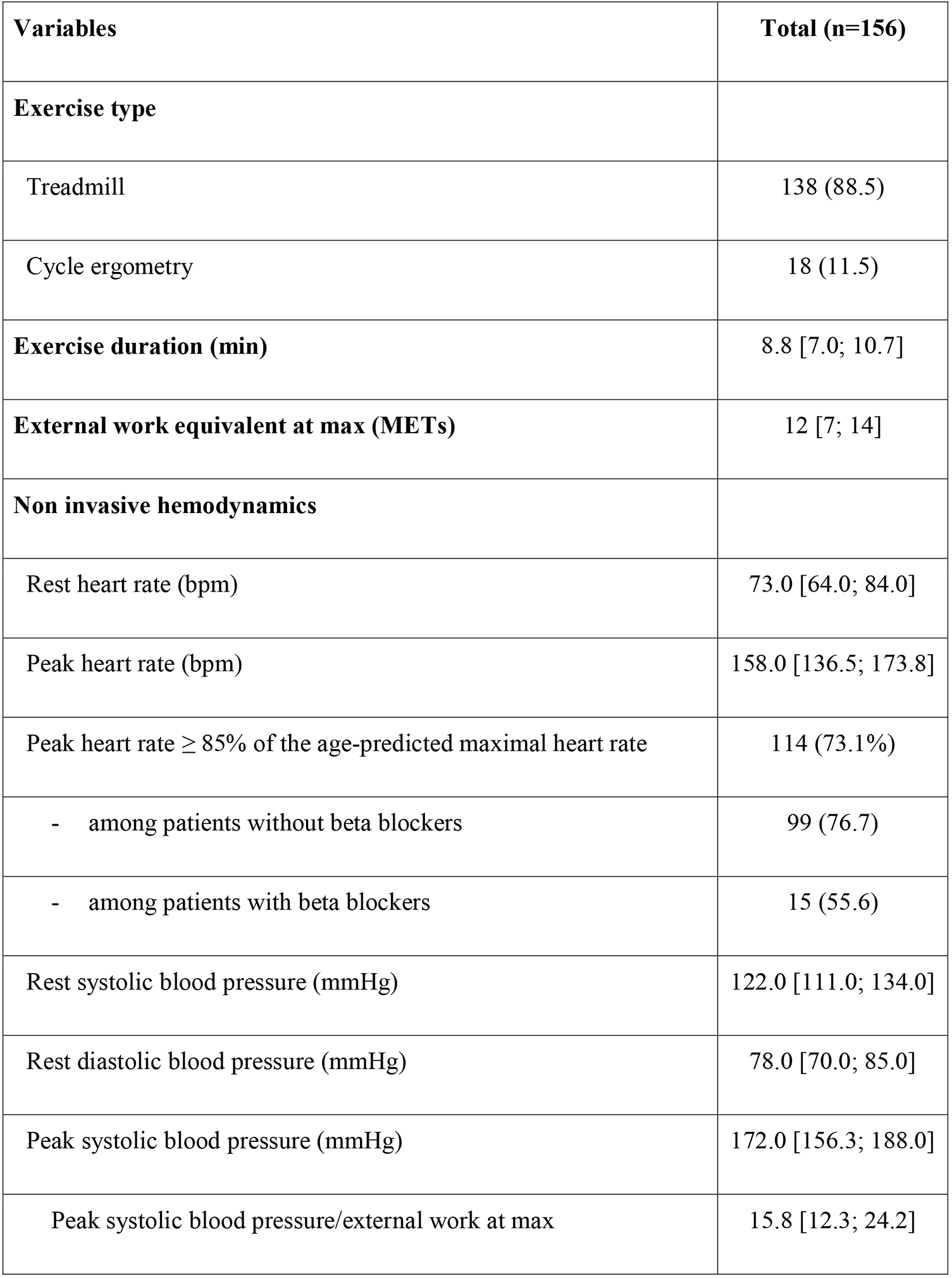
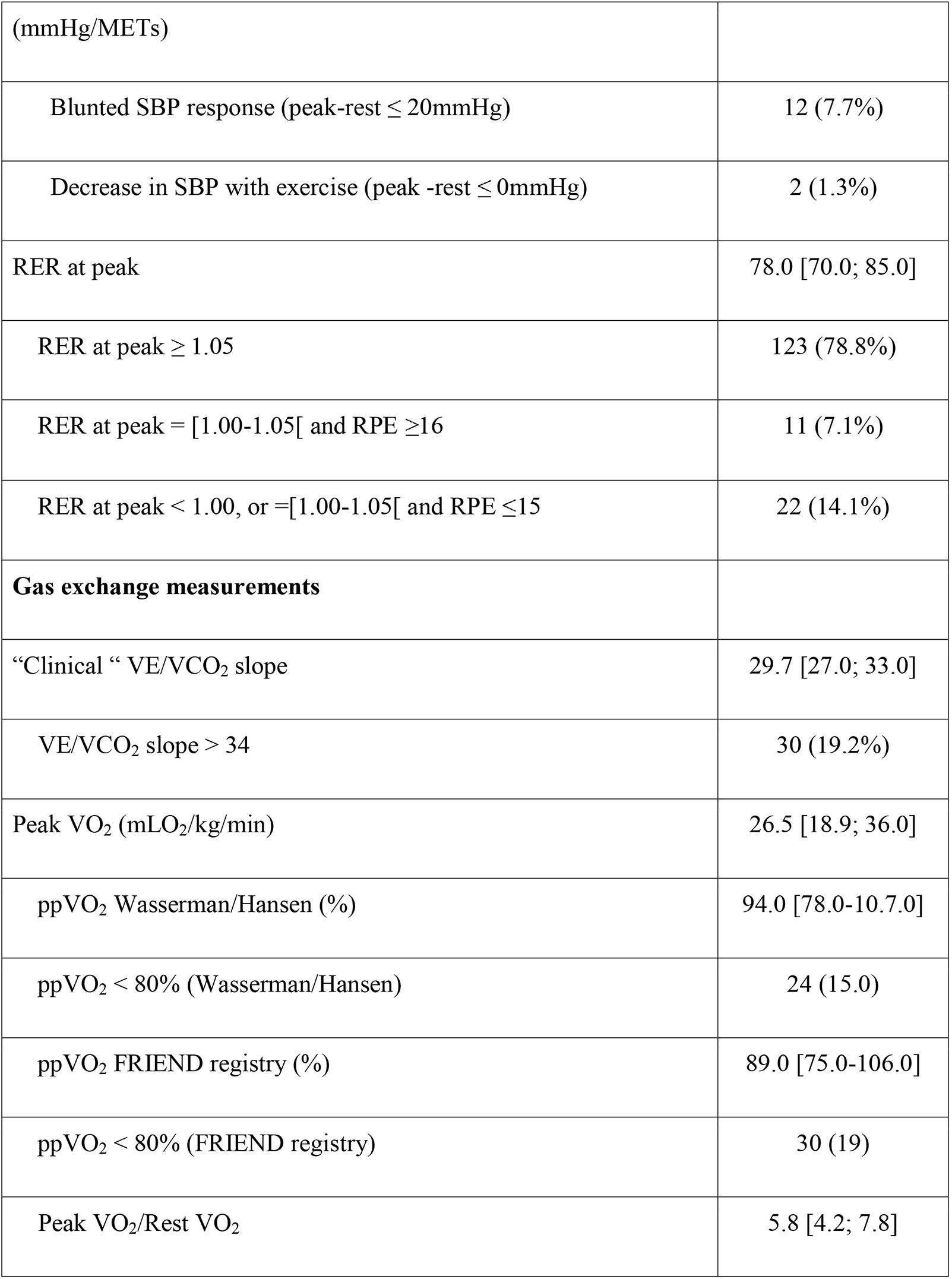
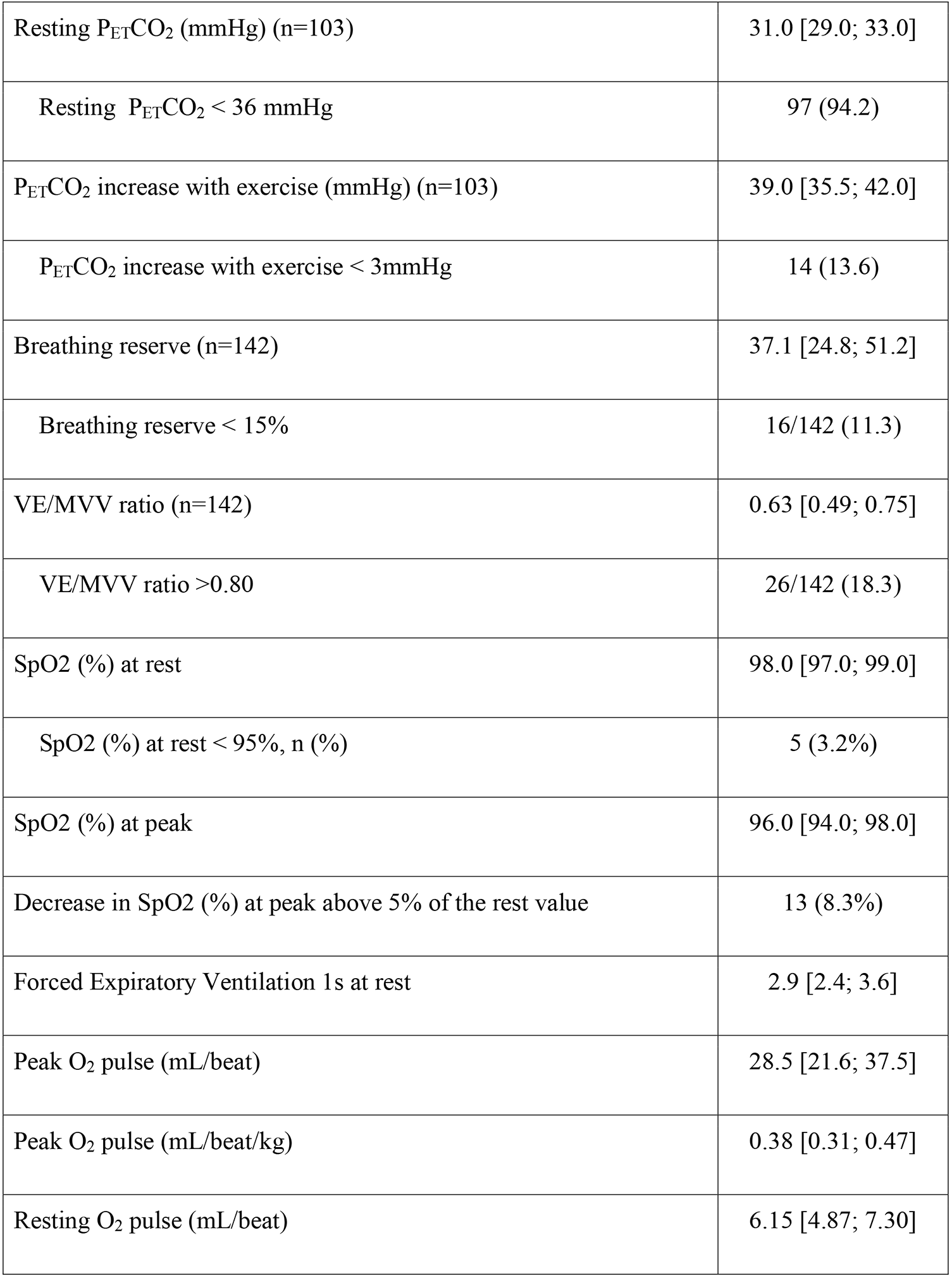
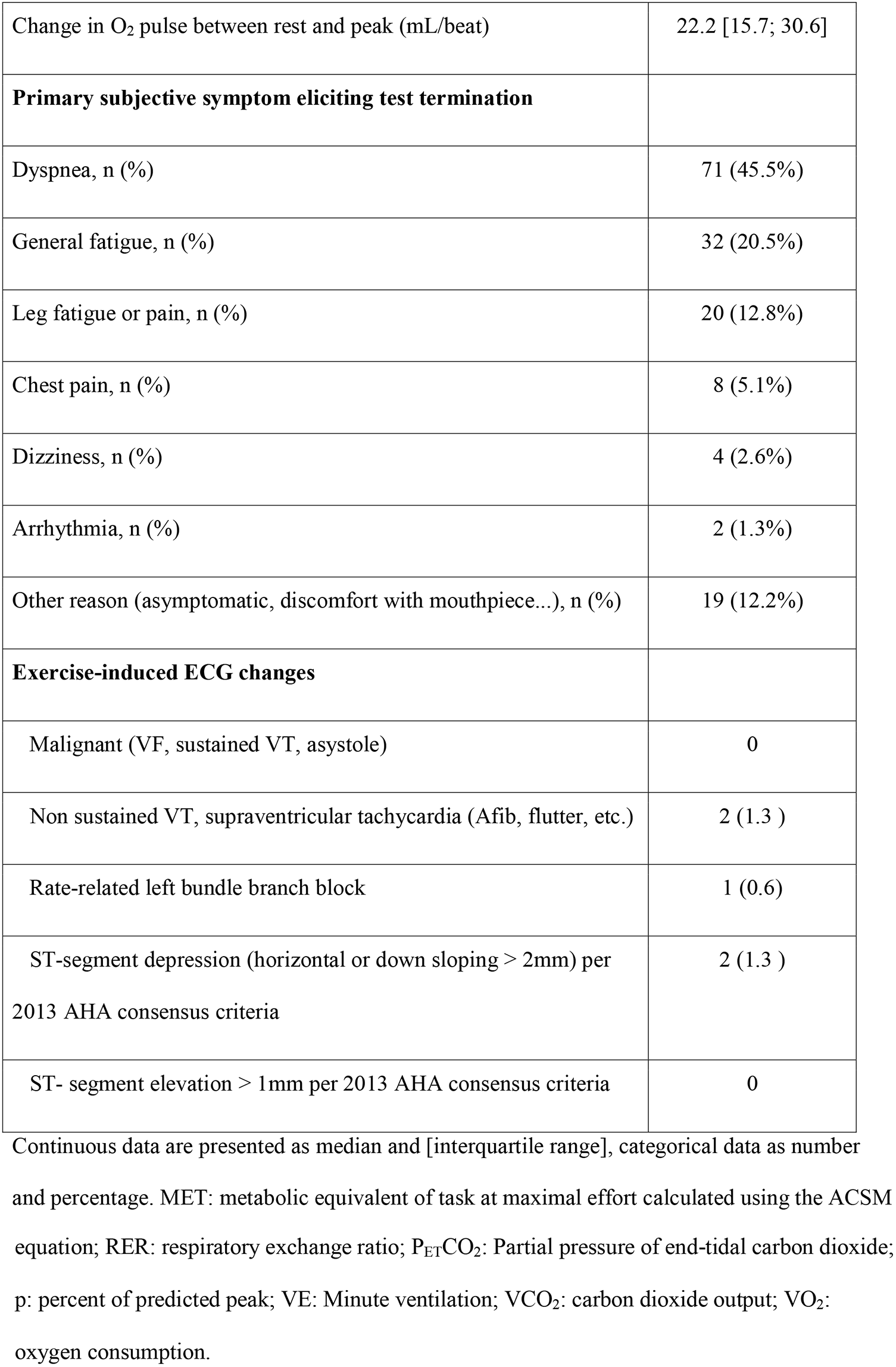
Cardiopulmonary exercise testing results of the total cohort (n=156).

**Figure 2.**
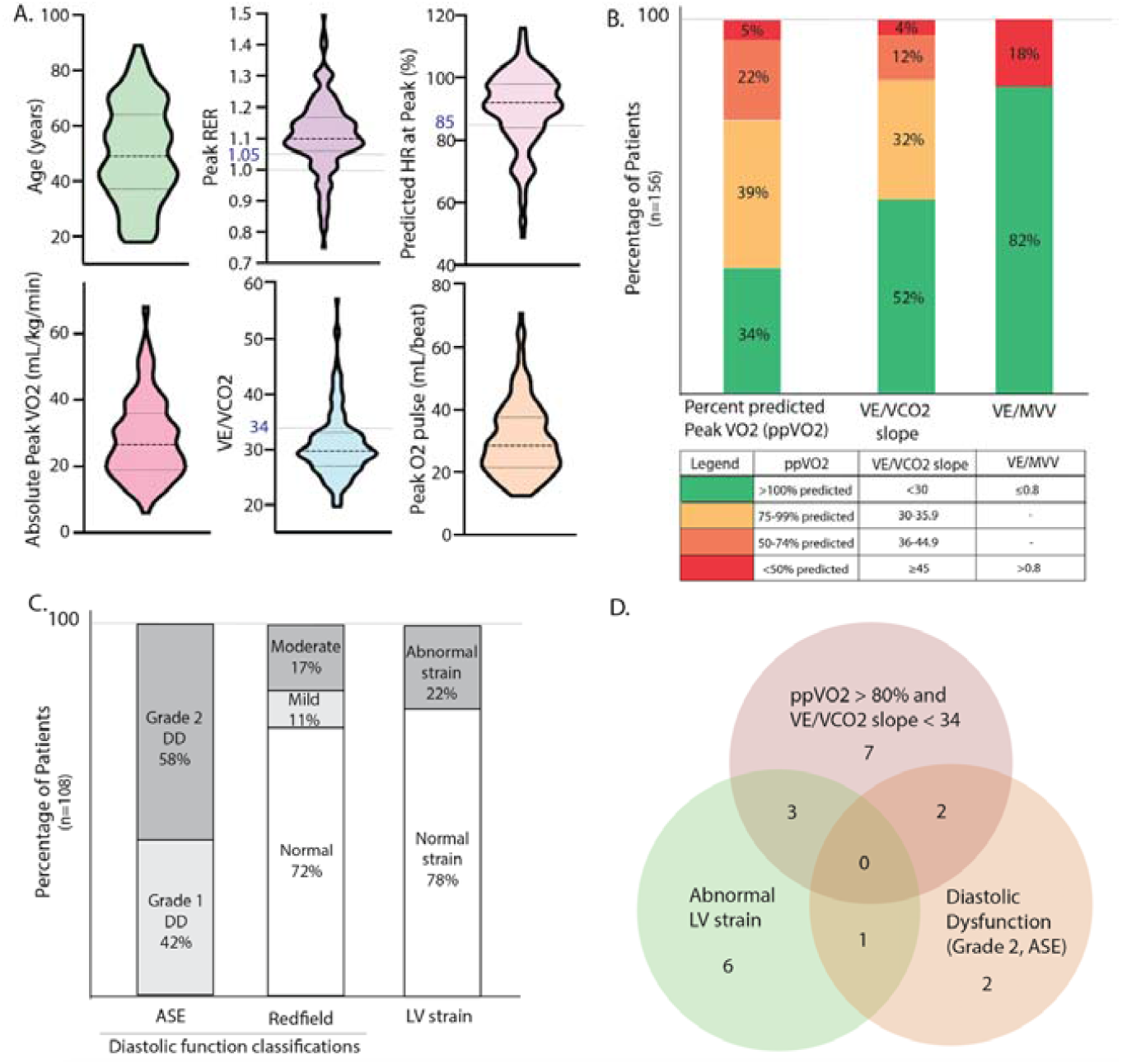
Violin plots of selected demographics and CPX data (A), prevalence of echocardiographic-based left ventricular dysfunction (B) and prevalence of diastolic dysfunction or abnormal strain in those with normal CPX data (C). Panel A: Thick dotted line represents the median, thin dotted lines represent the 25th and 75th percentile respectively. Panel B: Distribution of patients based on primary CPX variables from the 2016 guidelines by Guazzi et al (3). Panel C: Prevalence of diastolic dysfunction based on the American Society of Echocardiography (ASE) classification or the classification published by Redfield and prevalence of abnormal left ventricular (LV) global longitudinal strain using gender-specific normative values adapted from two papers by Kuznetzova (greater than -17.4% in men and - 18.8% in women) (12, 13, 15). Panel D: Prevalence of Grade 2 diastolic dysfunction based on ASE classification and/or abnormal gender-specific LV global longitudinal strain as in Panel C.

### Echocardiographic phenotyping

One-hundred and twenty-two patients had a stress and/or complete resting echocardiogram within 6 months of the date of CPX, including 118 in our institution. Among them, 87 patients had a stress echocardiogram with complete assessment of resting LV diastology, 21 had only resting echocardiogram with complete assessment of LV diastology, and 10 patients had a stress echocardiogram (without assessment of resting LV diastology).

Among the 97 patients with stress echocardiogram available for review, two patients (2.1%) presented with wall motion abnormalities during peak exercise/early recovery or late recovery in the territory of the left anterior descending coronary artery, which corresponded to coronary artery stenosis less than 50% of the lumen as assessed by coronary angiography. One patient had elevated RVSP (> 50mmHg) at peak exercise.

The baseline clinical characteristics of the 108 patients with complete resting LV diastology available for re-interpretation are presented in **Supplementary table S2**. Median age was 48 [36-61] years with 35% being male. There was no significant difference in the prevalence of cardiovascular risk factors between these 108 patients and the total cohort. **Table 3** summarizes their echocardiographic results. All patients had an LV ejection fraction above 50%. **Figure 2C** highlights the proportion of patients with underlying diastolic dysfunction based on three criteria [12-15].

**Table 3.** Resting echocardiographic findings of the subgroup with complete echocardiograms (n=108) for assessment of subclinical left ventricular (LV) dysfunction.

| **Variables** | **n=108** |
| --- | --- |
| LV end-diastolic internal diameter (cm) | 4.5 [4.2; 4.9] |
| LV mass index (g/m<sup>2</sup>) | 61.3 [50.1; 70.6] |
| LV geometry |  |
| Normal, % | 76 (70.4%) |
| Concentric remodeling, % | 31 (28.7%) |
| Concentric hypertrophy, % | 0 |
| Eccentric hypertrophy, % | 1 (0.9%) |
| LV ejection fraction (%) | 62 [58.8; 66.6] |
| LV global longitudinal strain (%) | -19.8 [-18.4; -22.2] |
| Left atrial maximal volume index (mL/m<sup>2</sup>) | 20.7 [15.4; 26.1] |
| Left atrial maximal volume index > 34.0 mL/m<sup>2</sup> | 8 (7.4%) |
| Left atrial emptying fraction (%) | 61.3 [55.3; 67.2] |
| Left atrial reservoir strain (%) | -24.8 [-22.3; -28.6] |
| Diastolic dysfunction per ASE guidelines, % | 50 (46.3%) |
| Grade 1, % | 21 (42%) |
| Grade 2, % | 29 (58%) |
| Average E/e' > 14 | 7 (6.5%) |
| Pulmonary vein S/D ratio >1, % | 62/88 (7%) |
| TR Vmax > 2.8m/s | 4 (3.7%) |
| RVSP (mmHg) at rest, n=63 | 24.2 [20.8; 30.0] |
| RV basal diameter, cm | 3.2 [2.9; 3.6] |
| RV mid diameter, cm | 2.3 [1.9; 2.6] |
| Tricuspid annular plane systolic excursion TAPSE, cm* | 2.4 [2.1; 2.7] |

\*All patients had a TAPSE > 1.7cm.

### Clinical diagnosis

Among the total cohort with unexplained dyspnea referred for CPX (n=156), 125 patients had a documented follow-up visit at our institution and affiliated institutions. One patient died of septic arthritis during follow-up. The clinical diagnoses are presented in **Supplementary Table S3**. During follow-up, the clinical diagnosis was attributed to cardiovascular etiology in 29 patients (18.6%, including heart failure, coronary artery disease, arrhythmia, moderate mitral regurgitation, hypertension), pulmonary etiology in 13 patients (8.3%, including obstructive lung disease, reactive airways disease and restrictive lung disease), pulmonary hypertension in 5 patients (3.2%), deconditioning in 15 patients (9.6%) and other conditions in 15 patients (including anxiety, vocal cord dysfunction, post-viral bronchial hyperreactivity, gastroesophageal reflux disease, malignancy, costochondritis, chronic fatigue syndrome, neuromuscular disease, medication-related). The dyspnea remained unexplained in 55 patients (35.3%). Among them, 31 patients had complete echocardiographic-based LV diastology for review: 12 had ppVO2 > 80 % and VE/VCO2 slope < 34, 5 patients had ASE Grade 2 diastolic dysfunction at rest and 10 had abnormal LV strain. When further analyzing early markers of LV subclinical dysfunction overlap with CPX results (**Figure 2D**), we identified 3 patients with ppVO2 > 80%, VE/VCO2 slope < 34 and abnormal strain, and 2 patients with ppVO2 > 80%, VE/VCO2 slope < 34 and Grade 2 diastolic dysfunction.

## DISCUSSION

Our study highlights the increasing number of patients with unexplained dyspnea referred for CPX. Subsequent analysis of the diagnostic work-up revealed marked heterogeneity. CPX analysis enabled identification of the etiology of dyspnea in a majority of patients. Assessment of early markers of LV dysfunction was incremental to CPX in identifying the cause of dyspnea. CPX is an under-utilized diagnostic tool that allows differentiation between pulmonary, cardiac and peripheral causes of exertional dyspnea through analysis of gas exchange and hemodynamic patterns [16 - 18]. CPX utility in HFrEF and unexplained exertional dyspnea is well established [19]. A landmark study by Mancini et al. in 1991 established peak VO2 as a valuable prognostic indicator in HFrEF and later, VE/VCO2 slope emerged as a complementary prognostic indicator [8, 20]. Exercise oscillatory ventilation and PET-CO2 have also emerged as newer prognostic markers in HFrEF [21]. Similarly, in HFpEF, the VE/VCO2 slope and exercise oscillatory ventilation have been found to have similar prognostic value as in HFrEF but further studies are needed to establish their true clinical significance [2]. Of note, Nedeljkovic et al. explored the utility of combining CPX with stress echocardiography in 87 patients with unexplained exertional dyspnea and hypertension with normal resting echocardiography results [22]. In this group, 9.2% had an E/e’ value greater than 15 which was reinforced by additional CPX abnormalities such as a higher VE/VCO2 slope and together, these findings were consistent with early HFpEF. Meanwhile, in unexplained exertional dyspnea, CPX and combined stress echo aim to reproduce the patient’s symptoms to identify the underlying cause [23].

Among the multiple US or European consensus statements proposing guidelines for CPX interpretation, Guazzi et al. succinctly summarized the diagnostic stratification for patients with heart failure, confirmed or suspected hypertrophic cardiomyopathy, suspected or confirmed pulmonary arterial hypertension, chronic obstructive pulmonary disease, interstitial lung disease, myocardial ischemia, mitochondrial myopathies and unexplained exertional dyspnea [3]. For patients with unexplained exertional dyspnea, current recommendations focus on the primary variables including VE/VCO2 slope, percent-predicted peak VO2, PETCO2 and VE/MVV. Other standard variables include PFTs, pulse oximetry, ECG and hemodynamic monitoring. Transitioning from the green to red zone indicates a decline in functional capacity secondary to exertional dyspnea. Based on the abnormal variables detected during CPX, this diagnostic stratification chart can be used to help elucidate the underlying etiology for an individual’s exertional dyspnea.

### Clinical Implications

The heterogeneity in diagnostic work-up for patients with unexplained dyspnea highlights the need for a standardized workflow to minimize delays in diagnosis and medical care. A dyspnea clinic, as implemented in other centers, offers a multi-disciplinary and symptom-based diagnostic approach to promote early intervention and treatment in patients with unexplained dyspnea [1]. A dyspnea clinic staffed by experts in Cardiology, Pulmonology and Exercise Physiology would enable streamlined and comprehensive evaluations of patients referred with unexplained dyspnea. The utility of advanced CPX testing in this setting is magnified as it facilitates early interpretation of results and communication with the primary physician who orders the test. In our study, there were many cases of patients referred for CPX without a subsequent follow-up visit at our institution to discuss the results or delayed follow-up visits pending CPX result interpretation as currently. As CPX remains an advanced diagnostic test requiring expert interpretation, we believe that an integrated dyspnea clinic would consolidate its value in complex patient populations. Moreover, targeted interventions such as prescribed physical activity, rehabilitation and exercise programs can be tailored based on CPX findings for individual patients [16].

### Study Limitations

The first limitation of our study was the retrospective design based on chart review, as a result of which some measured variables were not available for all patients. To overcome this, chart review encompassed thorough searches of outside records via the Care Everywhere feature on EPIC electronic health record software. However, a proportion of patients still do not have diastology data as part of their echocardiographic results which further reflects the heterogeneity in data collection. In addition, LV diastology assessment at peak exercise was not performed in our cohort, precluding identification of early exercise-induced diastolic dysfunction. There is now strong evidence supporting measurement of the E/e’ ratio at peak exercise (or early recovery), as peak elevated values are more predictive of elevated LV filling pressures-induced with exercise than resting values [3, 24 - 26]. Our study highlights the importance of peak diastology assessment in clinical practice for patients with unexplained dyspnea, which has since been routinely implemented in our practice. Lastly, our study was limited to the assessment of patients able to perform exercise testing, and the significance of unexplained dyspnea precluding exercise testing was not explored.

In conclusion, CPX is increasingly being used to investigate unexplained dyspnea, as it can identify the etiology of dyspnea in a majority of patients. The diagnostic work-up of patients with unexplained dyspnea remains markedly heterogeneous. Assessment of early markers of LV dysfunction is incremental to CPX in patients with dyspnea. These results highlight the need for a more standardized diagnostic workflow and the value of an integrated dyspnea clinic in facilitating early diagnosis and treatment.

## Data Availability

All data produced in the present study are available upon reasonable request to the authors.

## AUTHOR CONTRIBUTIONS

Pooja Nair, Jeffrey W Christle and Myriam Amsallem were involved in the conception and design of the work, data gathering, data analysis and interpretation, writing and critical revision of the article. Francois Haddad was involved in conception and design, data analysis and interpretation, writing and critical revision. Constance Verdonk, Nicholas Cauwenberghs, Yair Blumberg, Laura S Sasportas, Roham Zamanian, Euan Ashley, Tatiana Kuznetsova and Jonathan Myers contributed to data interpretation article writing and critical revision. All authors read and approved the final version of the manuscript.

## ACKNOWLEDGMENTS

The authors would like to thank Stanford Cardiovascular Institute and the Vera Moulton Wall Center at Stanford for Pulmonary Vascular Disease for their support.

